# Matters of the Heart and Soul: The Mixed Impact of Religiosity and Spirituality on the Cardiovascular Risk Profile Over Time

**DOI:** 10.64898/2026.09.14.26362903

**Authors:** Vinicius Vieira Neves, Cesar de Oliveira, LaPrincess C. Brewer, Ione Jayce Ceola Schneider, Andrew Steptoe

## Abstract

**Aims:** Religiosity and spirituality (R/S) are generally associated with a more favourable cardiovascular risk profile, but longitudinal evidence is limited. We examined whether multidimensional R/S predicts its future behavioural and biological components.

**Methods:** This prospective cohort study analysed 6,844 adults aged ≥50 (56% women; mean R/S score 2.1; 20% weekly attenders; 81% Christian) from waves 4–7 of the English Longitudinal Study of Ageing (ELSA). Behavioural outcomes included smoking, exercise, alcohol intake, and fruit and vegetable consumption; biological outcomes included blood pressure (BP), haemoglobin A1c (HbA1c), and the inflammatory risk markers C-reactive protein (CRP) and fibrinogen. R/S was assessed with four items from the Santa Clara Strength of Religious Faith Questionnaire. Hierarchical linear and logistic regressions adjusted for age, sex, wealth, education and ethnicity; biomarker models also adjusted for health behaviours and body mass index.

**Results:** Greater spirituality (*B*=−0.019; 95%CI[−0.036,−0.003]), daily prayer/meditation (*B*=−0.017; 95%CI[−0.032,−0.002]), participation in organized religion (*B*=−0.017; 95%CI[−0.032,−0.002]), and importance of religious faith (*B*=−0.017; 95%CI[−0.033,−0.002]) were independently associated with lower fibrinogen levels. Daily prayer/meditation also independently predicted higher fruit and vegetable intake (*B*=0.004; 95%CI[0.000,0.008]). However, frequent attendance (OR=0.846; 95%CI[0.730,0.982]), the significance of faith (OR=0.935; 95%CI[0.879,0.994]), and religious purpose (OR=0.939; 95%CI[0.884,0.997]) independently reduced the odds of meeting exercise recommendations. Similarly, frequent attendance was associated with higher HbA1c (*B*=0.002; 95%CI[0.000,0.005]). There were no independent associations with CRP or blood pressure.

**Conclusion:** R/S showed mixed prospective associations with the cardiovascular risk profile: favourable for fibrinogen and fruit and vegetable intake, unfavourable for physical activity and HbA1c.

**LAY SUMMARY:** We examined whether religious and spiritual involvement was related to later heart-related health behaviours and blood measurements in more than 6,800 English adults aged 50 and over. The associations ran in both directions, and the study cannot show that religion or spirituality causes these changes.

- Stronger religious and spiritual involvement was associated with lower fibrinogen, a blood protein linked to clotting.
- Daily prayer or meditation was associated with eating more fruit and vegetables.
- Regular attendance at religious services was associated with less physical activity and higher blood sugar.

GRAPHICAL ABSTRACT

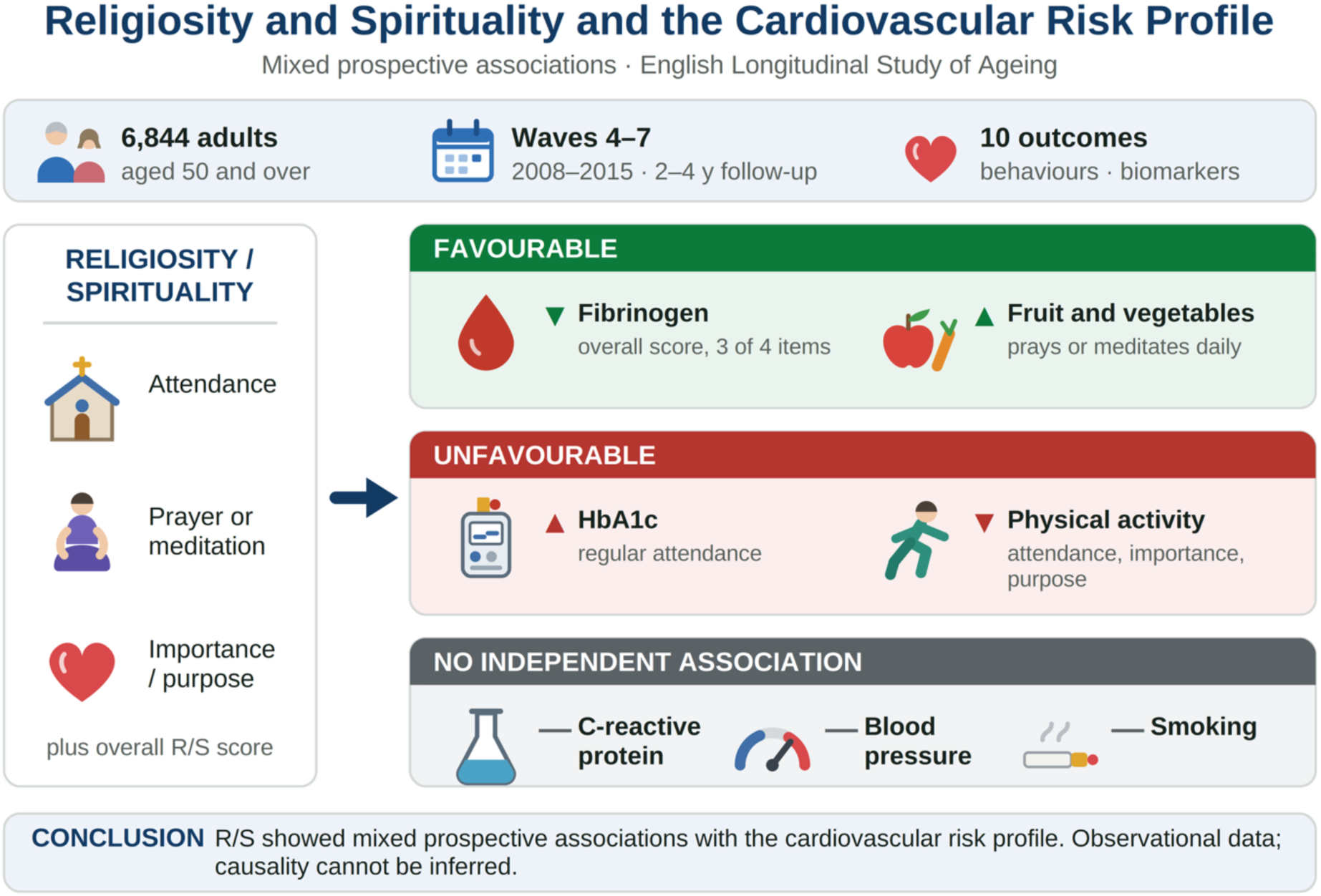

## INTRODUCTION

There is growing interest in the link between religiosity and spirituality (R/S) and health outcomes. Religiosity reflects how much one values and engages with one’s religion,^1^ whereas spirituality is defined as ‘an inherent aspect of human experience that arises from or leads to the search for purpose in life, and the connection with self, others, and the transcendent’.^1–3^ Initial research has consistently shown lower cardiovascular disease (CVD) mortality^1,4^ among those with higher R/S levels. However, more recent research presents somewhat contradictory results.^5^ For example, while frequent religious attendance predicted lower CVD mortality in an eight-year follow-up of the Nurses’ Health Study,^6^ analyses from the Third National Health and Nutrition Examination Survey (NHANES III) did not reach statistical significance.^7^ Additionally, non-organizational R/S, which includes engaging in R/S activities such as praying, meditating, or reading religious texts,^8^ was associated with a higher risk of CVD in a prospective analysis of the Women’s Health Initiative study.^9^ If these conflicting findings are genuine, they could be explained by the relationship between different aspects of R/S and behavioural or biological risk factors for CVD.

Several studies have examined the relationship between R/S and these behaviours. Koenig and colleagues estimate that 35% of the effects of R/S on CVD arise from behavioural mechanisms.^1^ Their systematic review of pre-2010 literature found that R/S was associated with protective effects against substance use and physical inactivity in most high-quality studies.^8^ Another systematic review suggests that R/S may increase fruit and vegetable consumption.^10^ However, most were cross-sectional and did not fully adjust for confounders such as income and ethnicity, limiting the ability to establish temporal relationships. Longitudinal studies with appropriate adjustments could address these gaps; to our knowledge, no prospective study has yet examined R/S and fruit and vegetable intake.

Regarding substance use, longitudinal studies primarily focus on American community-based cohorts. According to a 2022 evidence statement produced by a Delphi panel of experts, based on a systematic review of high-quality prospective studies,^5^ frequent religious attendance is associated with lower rates of smoking and drinking.

Considering evidence on healthier lifestyle behaviours, and even before the publication of the 2022 review, the Brazilian Society of Cardiology was the first to incorporate spirituality into cardiovascular prevention guidelines, emphasizing its importance for primary prevention strategies.^3^

Furthermore, R/S may influence CVD through direct physiological pathways. Emerging research independently links spiritual well-being, particularly inner peace and meaning, to the maintenance of endothelial function.^11^ A meta-analysis found that both organizational (e.g., service attendance) and non-organizational religiosity were associated with favourable biological measures, including blood pressure and C-reactive protein (CRP).^12^ However, findings from individual longitudinal studies are mixed: the Black Women’s Health Study reported that religious coping reduced incident hypertension,^13^ while the CARDIA Study found religious involvement increased incident hypertension before adjustment.^14^ Some population-based studies found no association between CRP and R/S.^7,15^

Another inflammatory marker, fibrinogen, was inversely correlated with religious attendance in some studies^7,16^ but not in others.^15^ Finally, two longitudinal studies on glycaemic control yielded conflicting results: religiosity was associated with higher HbA1c in one diabetic cohort^17^ but showed no effect in another.^18^ These mixed findings may partly explain why R/S is linked to obesity despite reducing overall CVD risk.^1,14,19^

Given that previous studies mainly used a cross-sectional design, focused on attendance as the primary R/S measure, failed to address confounding variables thoroughly, and yielded some conflicting results, further longitudinal research is necessary. Additionally, to our knowledge, no such study has been conducted with an English sample. Therefore, this study aims to examine the prospective relationship between multidimensional R/S and the cardiovascular risk profile in England. We hypothesize that (1) higher R/S will independently predict healthier behaviours, and (2) elevated R/S will be independently linked to improved biological markers.

## METHODS

### Study Design

The present study conducts a secondary data analysis using the English Longitudinal Study of Ageing (ELSA) to investigate the relationship between R/S and cardiovascular risk. ELSA is a prospective cohort study designed to represent the national population aged 50 and over living in England.^20^ Participants were initially recruited through the Health Survey for England between 1998 and 2001. The first wave of data collection took place in 2002, gathering psychosocial, economic, and health-related information. Participants were followed up every two years to complete a computer-assisted personal interview and every four years for biological assessments conducted by a nurse. Since the initial wave, ten additional waves of data have been collected, all approved by the National Research Ethics Service. All participants provided written consent, and specific approval for the secondary analysis conducted in this study was not necessary.

### Participants and Procedures

R/S measures were collected in ELSA wave 5 (2010/11). Of the 10,274 individuals who participated in this wave, 7,903 had at least one R/S variable. Of these, 6,844 had complete baseline and follow-up data on at least one outcome variable, forming the sample included in the primary analysis. As there were no physical assessments in wave 5, values from wave 4 (2008/9) were used as the baseline for biological covariates and outcomes.

Baseline sociodemographic and behavioural data were collected during wave 5. Therefore, the baseline data (T0) comprised waves 4 and 5, whereas follow-up data were collected in wave 6 (2012/13) for biological outcomes and in wave 7 (2014/15) for behavioural outcomes (T1) (Supplemental Table 1).

### Measures

#### Covariates and sociodemographic

*Age*: Participants were categorized into four age groups: 52-59, 60-69, 70-79, and 80+.

*Wealth:* Considering financial wealth (e.g., savings), the value of any property (net of mortgage), net debt, business assets, and physical wealth (e.g., artwork), SES was calculated and divided into quintiles. The wealthiest are in the higher quintiles, while the less affluent are in the lower quintiles.^21^

*Education*: educational attainment has been categorized into five levels: (i) no qualification; (ii) up to O level; (iii) A level and foreign qualifications; (iv) higher education below degree level; and (v) degree.

*Sex*: participants were classified as ‘female’ and ‘male’.

*Ethnicity*: Participants self-reported their ethnicity at their Health Survey for England baseline interview, using ELSA’s categories: White; Asian or Asian British; Black, African, Caribbean or Black British; Mixed ethnic group; and any other ethnic group. Ethnicity was included as a covariate because religiosity, cardiovascular risk factors and their social determinants differ across ethnic groups in England. Because each group other than White comprised fewer than 80 participants, these were combined for analysis, following current guidance.^22,23^ No comparisons between individual groups were made.

*Body Mass Index (BMI):* Height and weight were measured during nurse visits to calculate BMI. Height was measured with a portable stadiometer to the nearest millimetre, and weight with a Tanita THD-305 electronic scale to the nearest 0.1 kg. BMI categories were normal weight (BMI<25), overweight (BMI 25-29.9), class I obesity (BMI 30-34.9), class II obesity (BMI 35-39.9), and class III obesity (BMI≥40).

*Health behaviours, including* smoking status, physical activity, alcohol consumption, and fruit and vegetable intake, served as both covariates and outcomes in this study. The next section will detail the data collection methods for these behaviours (see measures: outcomes).

### Exposures

*Religiosity/spirituality:* Four questions adapted from the Santa Clara Strength of Religious Faith Questionnaire^24^ were used to develop the self-report scale for this study. The final instrument uses a four-point Likert scale (ranging from 1 = ‘strongly disagree’ to 4 = ‘strongly agree’) and includes the following items: (i) Religious faith is essential to me; (ii) I pray or meditate daily; (iii) I seek my religion to find meaning and purpose in life; (iv) I consider myself active in organized religion. Final scores range from 1 to 4, with higher scores indicating greater spirituality or religiosity.

The three-dimensional categorization of religiousness^25^ is a common approach in spirituality and health research. Items from this study’s scale were categorized as follows: items “i” and “iii”—religious importance; item “ii”—non-organizational religiosity; and item “iv”—organizational religiosity.

*Religious Attendance:* participants’ responses to the question ‘About how often have you attended religious services during the past year?’ were classified as ‘regular’ for those attending once a week or more, and ‘not regular’ for those attending three times a month or less.

*Importance in everyday life:* Participants were asked to respond to the question, ‘How important is religion in your daily life?’, on a four-point Likert scale, with 1 indicating very important and 4 indicating not at all important.

### Outcomes

Biological data were collected during nurse visits, including blood samples and blood pressure measurements. Participants taking anticoagulant medication or diagnosed with a coagulation disorder were ineligible to provide blood samples. All blood sample analyses were conducted at the Royal Victoria Infirmary in Newcastle upon Tyne, UK. Further details are described elsewhere.^26^

#### Cardiovascular Risk Factors

*Blood Pressure:* Pregnancy was the only exclusion criterion for obtaining BP readings. Participants were instructed not to consume alcohol, smoke, eat, or engage in vigorous exercise for at least 30 minutes before measurement. Using the Omron HEM-907 monitor, nurses recorded participants’ BP three times, with a 1-minute interval between measurements. We calculated the mean of the three measurements for systolic blood pressure (SBP) and diastolic blood pressure (DBP).

*Glycaemic Control:* Glycated haemoglobin (HbA1c) levels were measured using a Tosoh G7 analyser and reported as percentages.

#### Inflammatory Risk Markers

*C-Reactive Protein:* This was assessed using the N Latex CRP mono immunoassay on the Behring Nephelometer II analyser. CRP values were treated as a dichotomous variable centred on the well-accepted cut-off of 3mg/L,^27^ and classified as ‘high’ when above this level and ‘low/normal’ when below.

*Fibrinogen:* Serum concentrations (g/L) were measured using a Clauss thrombin-clotting method variant on the Organon Teknika MDA180 analyser.

#### Health Behaviours

*Smoking status:* Participants were asked whether they had ever smoked, and those who responded positively were then asked whether they still smoked. We classified responses as ‘yes’ for current smokers and ‘no’ for those who had never smoked or were past smokers, defined as having stopped at any point before the wave 5 interview.

*Physical activity (PA):* Participants were asked how often they took part in ‘sports or activities’ that were vigorous, moderately energetic, or mildly energetic. They could respond with ‘hardly ever or never’, ‘one to three times a month’, ‘once a week’, or ‘more than once a week’. In this study, responses were categorized as engaging in moderate or intense PA at least once a week or less than once a week.

*Sedentary Behaviour:* This was also assessed using responses to the PA questions. The summary index of PA measures was previously explained in detail.^28^ Individuals who did not engage in any PA category (mild, moderate, or vigorous) at least once a week were classified as sedentary.

*Alcohol Consumption:* Respondents reported the number of pints of beer, glasses of wine and measures of spirits consumed in the past seven days; these were converted to British units (one unit=8 g alcohol), a standard epidemiological method.^29^

*Intake of Fruits and Vegetables:* Participants were asked one question about fruit consumption and another about vegetable consumption. The first was, ‘How many portions of fruit (of any kind) do you eat on a typical day?’, while the second was, ‘How many portions of vegetables (excluding potatoes) do you eat on a typical day?’. Further instructions defined a portion of fruit as ‘an apple or banana, a small bowl of grapes, or three tablespoons of tinned or stewed fruit. If you drink fruit juice, you can count one glass per day, but additional glasses of fruit juice do not count as extra portions.’ A serving of vegetables was defined as ‘three heaped tablespoons of green or root vegetables such as carrots, parsnips, spinach, small vegetables like peas, baked beans, or sweet corn, or a medium bowl of salad (lettuce, tomatoes, etc.)’. Subsequently, these portions were summed to create a variable representing the total number of servings of fruits and vegetables that participants typically consume daily.

### Statistical Analyses

Simple correlations, reported as Pearson’s correlation coefficient (*r*) and *p*-values, were used to examine baseline associations between exposures and between exposures and covariates.

Skewed and/or kurtotic data were addressed through log transformation to ensure normal distribution.

Prospective associations between R/S and outcomes were analysed using multiple linear regression for continuous variables (e.g., SBP, HbA1c, drinking) and logistic regression for binary variables (e.g., CRP, smoking). Variables were entered hierarchically into the regression models. Model 1 adjusted for baseline outcome values (Wave 4 for biomarkers; Wave 5 for behaviours) and included one R/S exposure. Subsequently, Model 2 included age, sex, ethnicity, wealth, and education in analyses involving both biomarkers and health behaviours as outcomes. When biomarkers were outcomes, Model 2 also controlled for smoking status, physical activity, alcohol consumption, and fruit and vegetable intake. Finally, Model 3 further included BMI in the assessment of biomarkers. This order of entry was chosen to estimate the coefficient for the association between spiritual exposure and the health-related variable before controlling for covariates.^30^ Analyses used complete cases due to loss to follow-up. Results are presented as odds ratios (ORs), 95% confidence intervals (CIs), and *p*-values for logistic regression analyses, and as unstandardized regression coefficients (*B*), 95% CIs, and *p*-values for linear regression analyses. Statistical significance was set at *p*<0.05.

Further analyses were conducted for each item on the R/S scale to determine which dimension of R/S was most strongly associated with the outcomes.

All analyses were performed using IBM SPSS Statistics version 26.

## RESULTS

### Descriptive Statistics

The baseline characteristics of the 6,844 participants are presented in Table 1. Most participants were aged 60-69 (41%), 56% were female, 97% were White, and 81% were Christian. Approximately 20% regularly attended church, mosque, synagogue, etc. The mean spirituality score was 2.1, and 61% did not consider religion important in their daily lives.

**Table 1.** Baseline Characteristics of study participants (*N* = 6844), English Longitudinal of Ageing (ELSA), 2008-2015.

| Variables | *n* (%) |
| --- | --- |
| **Demographic Characteristics** |  |
| **Age group** |  |
| 50-59 | 1540 (22.5) |
| 60-69 | 2847 (41.6) |
| 70-79 | 1831 (26.8) |
| 80+ years | 626 (9.1) |
| **Sex** |  |
| Female | 3818 (55.8) |
| Male | 3026 (44.2) |
| **Wealth** |  |
| Lowest quintile | 1056 (15.4) |
| 2nd quintile | 1318 (19.3) |
| 3rd quintile | 1343 (19.6) |
| 4th quintile | 1465 (21.4) |
| Highest quintile | 1565 (22.9) |
| **Education\*** |  |
| No qualifications | 1606 (23.5) |
| Up to O level | 1604 (23.4) |
| A level of foreign qualification | 1083 (15.8) |
| Higher education below degree | 1143 (16.7) |
| Degree | 1406 (20.5) |
| **Ethnicity** |  |
| Any other ethnic group | 26 (0.4) |
| Asian or Asian British | 79 (1.2) |
| Black, African, Caribbean or Black British | 45 (0.7) |
| Mixed ethnic group | 14 (0.2) |
| White | 6677 (97.6) |
| Not reported | 3 (<0.1) |
| **Religiosity/Spirituality** |  |
| **Religion** |  |
| Christian | 5571 (81.4) |
| Other | 130 (1.9) |
| None | 1112 (16.2) |
| **R/S score (range 1-4) (mean ± SD)** | 2.1 ± 0.9 |
| **Attendance** |  |
| Never or seldom | 5350 (78.2) |
| Nearly every week or more | 1354 (19.8) |
| **Importance in everyday life** |  |
| Not at all or not very important | 4215 (61.6) |
| Somewhat or very important | 2567 (37.5) |
| **Biological Characteristics** |  |
| **CRP** |  |
| > 3.0mg/L | 1502 (31.9) |
| < 3.0mg/L | 3197 (68.1) |
| **Systolic BP (mmHg) (mean ± SD)** | 132.4 ± 17.3 |
| **Diastolic BP (mmHg) (mean ± SD)** | 74.6 ± 10.5 |
| **Fibrinogen (g/L) (mean ± SD)** | 3.3 ± 0.5 |
| **HbA1c (%) (mean ± SD)** | 5.8 ± 0.6 |
| **Health Behaviours** |  |
| **Current Smoker Status** |  |
| Yes | 794 (11.6) |
| No | 6050 (88.4) |
| **Moderate or vigorous physical activity** |  |
| Less than once per week | 2419 (35.3) |
| At least once per week | 4425 (64.7) |
| **Sedentary** |  |
| Yes | 348 (5.1) |
| No | 6496 (94.9) |
| **Units of alcohol in past week (mean ± SD)** | 2.8 ± 6.1 |
| **Total fruits and vegetables portions per day (mean ± SD)** | 5.0 ± 2.4 |

SD = standard deviation; R/S = Religiosity/Spirituality; CRP = C-Reactive Protein; BP = Blood Pressure; HbA1c = Glycated Haemoglobin
\*Education is the highest qualification recorded in ELSA, derived from the Health Survey for England baseline and ELSA interviews: no qualifications; up to O level (NVQ1–2, CSE, or GCE O level/GCSE, usually taken at age 16); A level or foreign qualification (NVQ3 or GCE A level, usually taken at age 18, together with qualifications obtained outside the UK or otherwise unclassified); higher education below degree; degree (NVQ4–5, degree or equivalent).

The three R/S measures were strongly intercorrelated: spirituality score with importance of religion in daily life (*r*=0.74), spirituality score with religious attendance (*r*=0.60), and importance of religion in daily life with religious attendance (*r*=0.63); all *p*<0.001. Higher scores on all three R/S measures were associated with older age and female sex (all *p*<0.001). Scores were also higher among participants of Asian, Black, Mixed or any other ethnic group than among White participants (all *p*<0.001). Spirituality scores and the importance of religion in daily life were higher among participants with lower wealth and lower educational attainment, whereas religious attendance was more frequent among wealthier and more educated participants (Supplemental Table 2).

### R/S and Biological Data

Hierarchical longitudinal multiple regression analyses were conducted to examine associations between R/S exposure and biological data (Table 2) and health behaviours (Table 3). Higher R/S scores were independently associated with lower fibrinogen levels two years later (*B*=−0.019; 95%CI[−0.036,−0.003]; *p*=0.023). Greater regular attendance was independently associated with higher HbA1c levels two years later (*B*=0.002; 95%CI[0.000,0.005]; *p*=0.037). Importance in daily life was not significantly associated with any biomarker in model 3. However, although no longer significant after controlling for covariates, higher religiosity/spirituality (*B*=−0.358; *p*=0.007) and greater importance in daily life (*B*=−0.323; *p*=0.005) were significantly associated with lower DBP two years later.

**Table 2.**
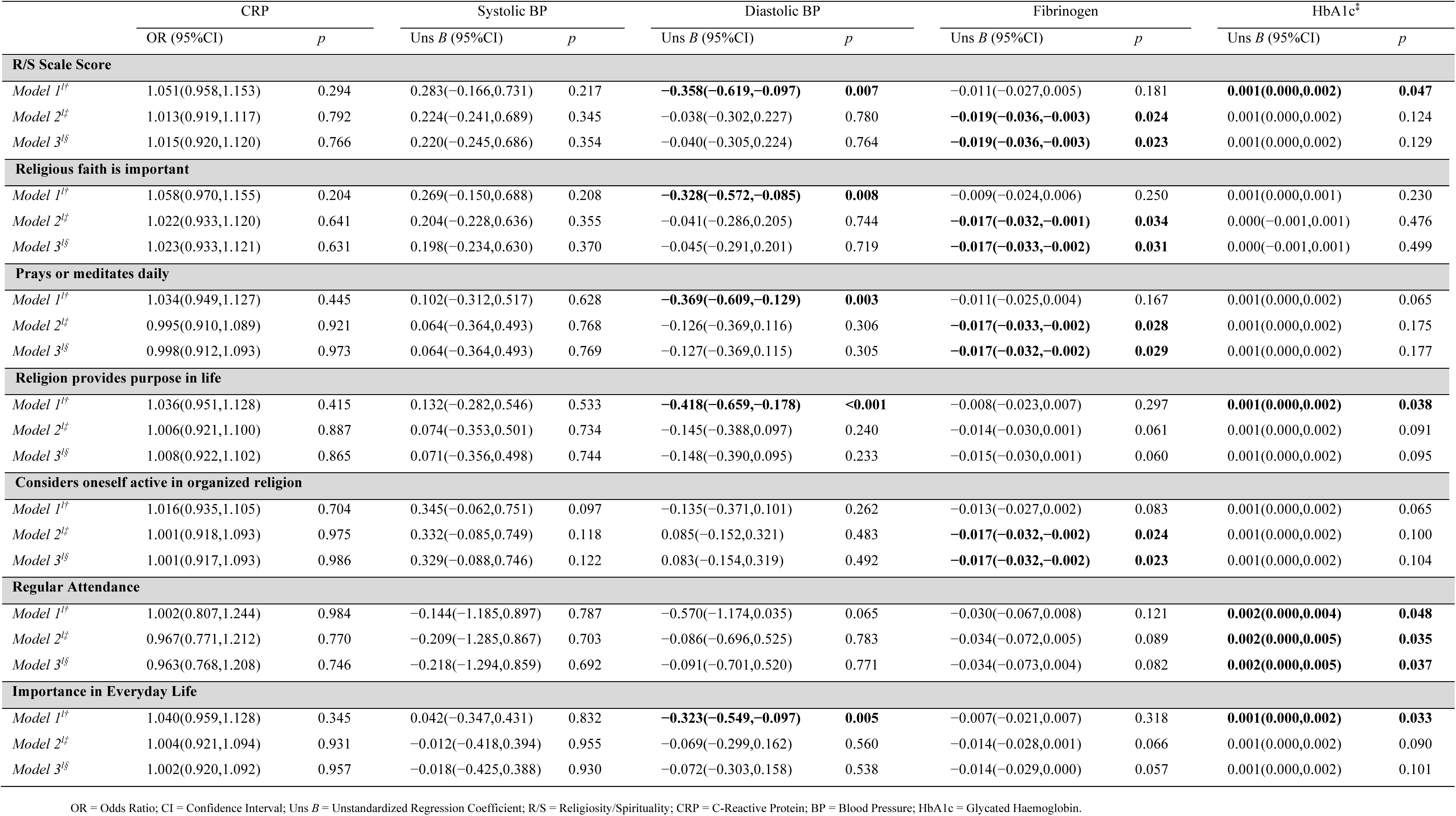
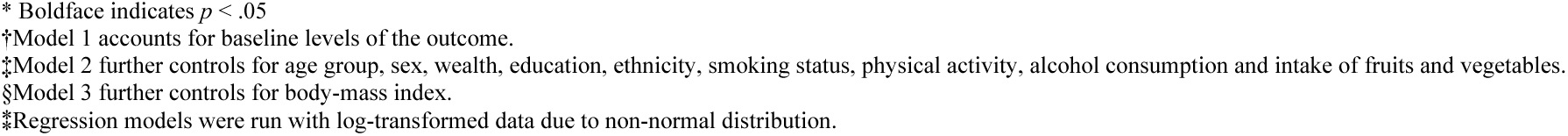
Prospective Associations Between R/S and Biomarkers, English Longitudinal of Ageing (ELSA), 2008-2015.

|  | CRP |  | Systolic BP |  | Diastolic BP |  | Fibrinogen |  | HbA1c<sup>‡</sup> |  |
| --- | --- | --- | --- | --- | --- | --- | --- | --- | --- | --- |
|  | OR (95%CI) | *p* | Uns *B* (95%CI) | *p* | Uns *B* (95%CI) | *p* | Uns *B* (95%CI) | *p* | Uns *B* (95%CI) | *p* |
| **R/S Scale Score** |  |  |  |  |  |  |  |  |  |  |
| *Model 1<sup>††</sup>* | 1.051(0.958,1.153) | 0.294 | 0.283(−0.166,0.731) | 0.217 | **−0.358(−0.619,−0.097)** | **0.007** | −0.011(−0.027,0.005) | 0.181 | **0.001(0.000,0.002)** | **0.047** |
| *Model 2<sup>‡‡</sup>* | 1.013(0.919,1.117) | 0.792 | 0.224(−0.241,0.689) | 0.345 | −0.038(−0.302,0.227) | 0.780 | **−0.019(−0.036,−0.003)** | **0.024** | 0.001(0.000,0.002) | 0.124 |
| *Model 3<sup>§§</sup>* | 1.015(0.920,1.120) | 0.766 | 0.220(−0.245,0.686) | 0.354 | −0.040(−0.305,0.224) | 0.764 | **−0.019(−0.036,−0.003)** | **0.023** | 0.001(0.000,0.002) | 0.129 |
| **Religious faith is important** |  |  |  |  |  |  |  |  |  |  |
| *Model 1<sup>††</sup>* | 1.058(0.970,1.155) | 0.204 | 0.269(−0.150,0.688) | 0.208 | **−0.328(−0.572,−0.085)** | **0.008** | −0.009(−0.024,0.006) | 0.250 | 0.001(0.000,0.001) | 0.230 |
| *Model 2<sup>‡‡</sup>* | 1.022(0.933,1.120) | 0.641 | 0.204(−0.228,0.636) | 0.355 | −0.041(−0.286,0.205) | 0.744 | **−0.017(−0.032,−0.001)** | **0.034** | 0.000(−0.001,0.001) | 0.476 |
| *Model 3<sup>§§</sup>* | 1.023(0.933,1.121) | 0.631 | 0.198(−0.234,0.630) | 0.370 | −0.045(−0.291,0.201) | 0.719 | **−0.017(−0.033,−0.002)** | **0.031** | 0.000(−0.001,0.001) | 0.499 |
| **Prays or meditates daily** |  |  |  |  |  |  |  |  |  |  |
| *Model 1<sup>††</sup>* | 1.034(0.949,1.127) | 0.445 | 0.102(−0.312,0.517) | 0.628 | **−0.369(−0.609,−0.129)** | **0.003** | −0.011(−0.025,0.004) | 0.167 | 0.001(0.000,0.002) | 0.065 |
| *Model 2<sup>‡‡</sup>* | 0.995(0.910,1.089) | 0.921 | 0.064(−0.364,0.493) | 0.768 | −0.126(−0.369,0.116) | 0.306 | **−0.017(−0.033,−0.002)** | **0.028** | 0.001(0.000,0.002) | 0.175 |
| *Model 3<sup>§§</sup>* | 0.998(0.912,1.093) | 0.973 | 0.064(−0.364,0.493) | 0.769 | −0.127(−0.369,0.115) | 0.305 | **−0.017(−0.032,−0.002)** | **0.029** | 0.001(0.000,0.002) | 0.177 |
| **Religion provides purpose in life** |  |  |  |  |  |  |  |  |  |  |
| *Model 1<sup>††</sup>* | 1.036(0.951,1.128) | 0.415 | 0.132(−0.282,0.546) | 0.533 | **−0.418(−0.659,−0.178)** | **<0.001** | −0.008(−0.023,0.007) | 0.297 | **0.001(0.000,0.002)** | **0.038** |
| *Model 2<sup>‡‡</sup>* | 1.006(0.921,1.100) | 0.887 | 0.074(−0.353,0.501) | 0.734 | −0.145(−0.388,0.097) | 0.240 | −0.014(−0.030,0.001) | 0.061 | 0.001(0.000,0.002) | 0.091 |
| *Model 3<sup>§§</sup>* | 1.008(0.922,1.102) | 0.865 | 0.071(−0.356,0.498) | 0.744 | −0.148(−0.390,0.095) | 0.233 | −0.015(−0.030,0.001) | 0.060 | 0.001(0.000,0.002) | 0.095 |
| **Considers oneself active in organized religion** |  |  |  |  |  |  |  |  |  |  |
| *Model 1<sup>††</sup>* | 1.016(0.935,1.105) | 0.704 | 0.345(−0.062,0.751) | 0.097 | −0.135(−0.371,0.101) | 0.262 | −0.013(−0.027,0.002) | 0.083 | 0.001(0.000,0.002) | 0.065 |
| *Model 2<sup>‡‡</sup>* | 1.001(0.918,1.093) | 0.975 | 0.332(−0.085,0.749) | 0.118 | 0.085(−0.152,0.321) | 0.483 | **−0.017(−0.032,−0.002)** | **0.024** | 0.001(0.000,0.002) | 0.100 |
| *Model 3<sup>§§</sup>* | 1.001(0.917,1.093) | 0.986 | 0.329(−0.088,0.746) | 0.122 | 0.083(−0.154,0.319) | 0.492 | **−0.017(−0.032,−0.002)** | **0.023** | 0.001(0.000,0.002) | 0.104 |
| **Regular Attendance** |  |  |  |  |  |  |  |  |  |  |
| *Model 1<sup>††</sup>* | 1.002(0.807,1.244) | 0.984 | −0.144(−1.185,0.897) | 0.787 | −0.570(−1.174,0.035) | 0.065 | −0.030(−0.067,0.008) | 0.121 | **0.002(0.000,0.004)** | **0.048** |
| *Model 2<sup>‡‡</sup>* | 0.967(0.771,1.212) | 0.770 | −0.209(−1.285,0.867) | 0.703 | −0.086(−0.696,0.525) | 0.783 | −0.034(−0.072,0.005) | 0.089 | **0.002(0.000,0.005)** | **0.035** |
| *Model 3<sup>§§</sup>* | 0.963(0.768,1.208) | 0.746 | −0.218(−1.294,0.859) | 0.692 | −0.091(−0.701,0.520) | 0.771 | −0.034(−0.073,0.004) | 0.082 | **0.002(0.000,0.005)** | **0.037** |
| **Importance in Everyday Life** |  |  |  |  |  |  |  |  |  |  |
| *Model 1<sup>††</sup>* | 1.040(0.959,1.128) | 0.345 | 0.042(−0.347,0.431) | 0.832 | **−0.323(−0.549,−0.097)** | **0.005** | −0.007(−0.021,0.007) | 0.318 | **0.001(0.000,0.002)** | **0.033** |
| *Model 2<sup>‡‡</sup>* | 1.004(0.921,1.094) | 0.931 | −0.012(−0.418,0.394) | 0.955 | −0.069(−0.299,0.162) | 0.560 | −0.014(−0.028,0.001) | 0.066 | 0.001(0.000,0.002) | 0.090 |
| *Model 3<sup>§§</sup>* | 1.002(0.920,1.092) | 0.957 | −0.018(−0.425,0.388) | 0.930 | −0.072(−0.303,0.158) | 0.538 | −0.014(−0.029,0.000) | 0.057 | 0.001(0.000,0.002) | 0.101 |

OR = Odds Ratio; CI = Confidence Interval; Uns *B* = Unstandardized Regression Coefficient; R/S = Religiosity/Spirituality; CRP = C-Reactive Protein; BP = Blood Pressure; HbA1c = Glycated Haemoglobin.
\* Boldface indicates $p < .05$
†Model 1 accounts for baseline levels of the outcome.
‡Model 2 further controls for age group, sex, wealth, education, ethnicity, smoking status, physical activity, alcohol consumption and intake of fruits and vegetables.
§Model 3 further controls for body-mass index.
\*Regression models were run with log-transformed data due to non-normal distribution.

**Table 3.** Prospective Associations Between R/S and Health Behaviours.

|  | Current Smoker |  | Moderate/Intense PA |  | Sedentary Behaviour |  | Alcohol Units<sup>§</sup> |  | Fru/Veg Intake<sup>§</sup> |  |
| --- | --- | --- | --- | --- | --- | --- | --- | --- | --- | --- |
|  | OR (95%CI) | *p* | OR (95%CI) | *p* | OR (95%CI) | *p* | Uns *B* (95%CI) | *p* | Uns *B* (95%CI) | *p* |
| **R/S Scale Score** |  |  |  |  |  |  |  |  |  |  |
| *Model 1*<sup>†</sup> | 1.033(0.875,1.218) | 0.702 | **0.849(0.799,0.901)** | **<0.001** | **1.201(1.078,1.338)** | **0.001** | **−0.016(−0.025,−0.007)** | **0.001** | 0.002(−0.003,0.006) | 0.479 |
| *Model 2*<sup>‡</sup> | 1.065(0.898,1.264) | 0.467 | 0.937(0.878,1.001) | 0.052 | 1.056(0.937,1.190) | 0.372 | −0.005(−0.014,0.004) | 0.239 | 0.002(−0.002,0.007) | 0.349 |
| **Religious faith is important** |  |  |  |  |  |  |  |  |  |  |
| *Model 1*<sup>†</sup> | 1.027(0.883,1.193) | 0.733 | **0.860(0.813,0.910)** | **<0.001** | **1.190(1.073,1.320)** | **0.001** | **−0.010(−0.018,−0.002)** | **0.019** | 0.001(−0.003,0.005) | 0.527 |
| *Model 2*<sup>‡</sup> | 1.060(0.907,1.238) | 0.463 | **0.935(0.879,0.994)** | **0.031** | 1.066(0.952,1.193) | 0.268 | −0.001(−0.009,0.008) | 0.888 | 0.002(−0.002,0.006) | 0.378 |
| **Prays or meditates daily** |  |  |  |  |  |  |  |  |  |  |
| *Model 1*<sup>†</sup> | 1.064(0.917,1.235) | 0.411 | **0.877(0.830,0.926)** | **<0.001** | **1.235(1.120,1.361)** | **<0.001** | **−0.016(−0.024,−0.007)** | **<0.001** | 0.003(−0.001,0.007) | 0.118 |
| *Model 2*<sup>‡</sup> | 1.087(0.932,1.268) | 0.287 | 0.962(0.906,1.022) | 0.209 | 1.099(0.988,1.223) | 0.082 | −0.006(−0.015,0.002) | 0.127 | **0.004(0.000,0.008)** | **0.049** |
| **Religion provides purpose in life** |  |  |  |  |  |  |  |  |  |  |
| *Model 1*<sup>†</sup> | 0.988(0.853,1.144) | 0.870 | **0.868(0.821,0.918)** | **<0.001** | **1.128(1.020,1.247)** | **0.019** | **−0.012(−0.021,−0.004)** | **0.003** | 0.001(−0.003,0.006) | 0.466 |
| *Model 2*<sup>‡</sup> | 1.023(0.879,1.191) | 0.770 | **0.939(0.884,0.997)** | **0.040** | 1.003(0.898,1.119) | 0.961 | −0.004(−0.012,0.004) | 0.360 | 0.002(−0.002,0.006) | 0.390 |
| **Considers oneself active in organized religion** |  |  |  |  |  |  |  |  |  |  |
| *Model 1*<sup>†</sup> | 0.954(0.817,1.114) | 0.549 | **0.909(0.861,0.960)** | **0.001** | 1.064(0.964,1.175) | 0.220 | **−0.010(−0.019,−0.002)** | **0.012** | 0.000(−0.004,0.004) | 0.957 |
| *Model 2*<sup>‡</sup> | 0.982(0.838,1.151) | 0.822 | 0.964(0.909,1.022) | 0.217 | 0.989(0.888,1.102) | 0.847 | −0.004(−0.012,0.004) | 0.300 | 0.000(−0.004,0.004) | 0.846 |
| **Regular Attendance** |  |  |  |  |  |  |  |  |  |  |
| *Model 1*<sup>†</sup> | 0.834(0.529,1.315) | 0.435 | **0.778(0.678,0.892)** | **<0.001** | 1.140(0.891,1.459) | 0.296 | **−0.032(−0.053,−0.012)** | **0.002** | 0.000(−0.011,0.010) | 0.939 |
| *Model 2*<sup>‡</sup> | 0.896(0.564,1.424) | 0.642 | **0.846(0.730,0.982)** | **0.027** | 1.004(0.770,1.309) | 0.977 | −0.019(−0.040,0.002) | 0.072 | −0.003(−0.014,0.007) | 0.527 |
| **Importance in Everyday Life** |  |  |  |  |  |  |  |  |  |  |
| *Model 1*<sup>†</sup> | 1.044(0.909,1.199) | 0.545 | **0.881(0.837,0.928)** | **<0.001** | **1.115(1.016,1.223)** | **0.022** | **−0.016(−0.024,−0.008)** | **<0.001** | 0.002(−0.002,0.006) | 0.338 |
| *Model 2*<sup>‡</sup> | 1.070(0.927,1.236) | 0.354 | 0.954(0.902,1.010) | 0.109 | 1.016(0.918,1.126) | 0.755 | −0.006(−0.014,0.002) | 0.141 | 0.002(−0.001,0.006) | 0.222 |

OR = Odds Ratio; CI = Confidence Interval; Uns *B* = Unstandardized Regression Coefficient; R/S = Religiosity/Spirituality; PA = Physical Activity; Fru/Veg = Fruits and Vegetables.\* Boldface indicates *p* < .05
†Model 1 accounts for baseline levels of the outcome.
‡Model 2 further controls for age group, sex, wealth, education and ethnicity.
§Regression models were run with log-transformed data due to non-normal distribution.

When analysing each of the four items comprising the R/S scale separately, we observed a significant negative association between praying or meditating daily and fibrinogen levels two years later (*B*=−0.017; 95%CI[−0.032,−0.002]; *p*=0.029) (Table 2).

This association was independent of age, ethnicity, sex, education, wealth, smoking status, physical activity, alcohol intake, fruit and vegetable consumption, and BMI. Similar negative independent relationships were found between considering oneself active in organized religion and fibrinogen (*B*=−0.017; 95%CI[−0.032,−0.002]; *p*=0.023), and between considering religious faith important and fibrinogen (*B*=−0.017; 95%CI[−0.033,−0.002]; *p*=0.031). However, the significant associations of religious faith being important, praying or meditating daily, and seeking religion to provide meaning with lower DBP after two years did not remain significant after adjusting for covariates. Covariates also explained the relationship between higher HbA1c and seeking religion to provide meaning. Finally, CRP and SBP remained unrelated to any R/S exposures.

### R/S and Health Behaviours

Individuals reporting higher baseline religious attendance had 15.4% lower odds of engaging in moderate– or high-intensity physical activity over the subsequent four years (OR=0.846; 95%CI[0.730,0.982]; *p*=0.027), independent of covariates. In the prospective analysis of Model 2, importance in daily life and R/S scores were not associated with any health behaviour. Although no longer significant after adjusting for covariates, higher religiosity/spirituality (OR=0.849; *p*<0.001) and greater importance in daily life (OR=0.881; *p*<0.001) were significantly associated with reduced odds of participating in moderate or intense physical activity four years later. Higher spirituality and importance were also significantly associated with increased odds of being sedentary, although this association was accounted for covariates. Finally, participants with higher levels of spirituality, who attended more religious institutions and reported greater importance, consumed significantly less alcohol after four years. However, this relationship was not significant after adjustments in Model 2.

Table 3 also summarizes the results of a multivariate analysis of the relationship between health behaviours and individual items on the R/S scale. The odds of engaging in moderate-to-intense physical activity were independently lower among individuals who reported that religious faith holds extreme importance for them (OR=0.935; 95%CI[0.879,0.994]; *p*=0.031) and among those seeking religion to provide meaning in their lives (OR=0.939; 95%CI[0.884,0.997]; *p*=0.040). However, individuals who reported daily prayer or meditation consumed significantly more fruits and vegetables over the following four years, even after adjusting for covariates (*B*=0.004; 95%CI[0.000,0.008]; *p*=0.049). Neither cigarette nor alcohol use nor sedentary behaviour was independently associated with R/S exposures. Nonetheless, all four items of the R/S scale were significantly associated with reduced alcohol intake after four years in model 1.

## DISCUSSION

Our findings suggest that higher R/S has mixed prospective associations with the cardiovascular risk profile. It was independently associated with lower fibrinogen, an inflammatory risk marker, and with greater fruit and vegetable intake. However, higher R/S was also independently associated with worse glycaemic control (higher HbA1c) and lower participation in moderate-to-vigorous exercise. These associations were independent of sex, age, wealth, education, and ethnicity; associations with HbA1c and fibrinogen also remained independent of smoking, physical activity, alcohol intake, fruit and vegetable intake, and BMI. Although our results indicate prospective links between higher R/S and more sedentary behaviour, lower alcohol intake, and reduced DBP, these were not independent of covariates. Finally, SBP, CRP and smoking status were not associated with dimensions of spirituality. Therefore, these results only partially support our hypotheses regarding R/S and the biological and behavioural components of the cardiovascular risk profile.

### R/S and Inflammatory Markers

Previous research linking R/S and inflammation only partly supports our findings. Some studies have examined the relationship between R/S and fibrinogen. Cross-sectionally, greater religious attendance was associated with lower fibrinogen in NHANES III and the MacArthur Successful Aging Study.^7,16^ However, prospective population-based studies from the United States (US) and Taiwan found no such association.^15^ This inconsistency may arise from differing definitions of frequent attendance across studies. We regarded ‘frequent’ as weekly, whereas others defined it as monthly.^7,15,16^

Additionally, two studies assessed attendance as part of a social integration index rather than directly.^15,16^ Regarding organizational religiosity, attendance was not linked to lower fibrinogen levels; conversely, ‘considering oneself active’ was a significant independent correlate. Future research could explore whether this relationship is mediated by self-reported attendance, participation in other R/S-organized activities, or perceptions of engagement. In our study, lower fibrinogen was also independently associated with the overall R/S score, considering religious faith essential, and daily prayer or meditation, spanning all three dimensions of religiousness. To our knowledge, this is the first study to examine these associations.

Another inflammatory marker associated with increased risk of CVD is CRP,^31^ and its link with spirituality has also been explored.^12^ We found no correlation between R/S and CRP, even prior adjustment. Two of four longitudinal analyses from population-based studies support our findings.^7,15^ Although the remaining analyses, based on the Health and Retirement Study (HRS)^32^ and the National Social Life, Health and Aging Project (NSHAP)^33^ cohorts of US older adults, identified independent inverse relationships between R/S and CRP, their results are not directly contradictory to ours.

The HRS, for example, found that religious beliefs and values predicted lower CRP levels after eight years of follow-up.^32^ This aspect of R/S was not assessed in our study, and their follow-up period was four times longer than ours. Given that the effects of R/S on biological markers may take time to emerge, a longer follow-up may reveal significant associations in our cohort. Additionally, in the HRS study, religious service attendance was not significantly associated with lower CRP, consistent with our findings.

Meanwhile, the NSHAP findings were significant only among Black participants, whereas our sample is 97% White, making direct comparisons between studies inappropriate.^33^ Prior research suggests that African Americans with high racial centrality and perceived racism tend to exhibit elevated CRP levels; however, when religious intensity is high, CRP levels appear to decrease.^34^ Perhaps religiosity buffers the inflammatory effects of systemic racism. Therefore, longitudinal data are broadly consistent with our results, showing no short-term association between religiosity and CRP in predominantly White samples.

### R/S and Physical Activity

Regarding cardiovascular risk factors, lifestyle can indirectly link R/S to health.^1^ Unexpectedly, our results indicate an independent decrease in moderate/vigorous physical activity (MVPA) at follow-up among individuals with higher R/S orientations. Only 2 of 21 higher-quality studies in a review by Koenig et al. reported the same inverse relationship.^8^ Longitudinal data examining this relationship remain limited. A recent prospective analysis of the HRS cohort showed that higher religious service attendance predicted future increases in exercise levels.^35^ Similarly, the SHARE study of 23,864 adults aged 50+ in nine European countries found that religious organizational involvement was associated with lower odds of physical inactivity over 11 years of follow-up.^36^

Several factors may explain the differences between these findings and ours. First, measurement thresholds differed: the HRS classified participants as “at least some exercisers” if they engaged in moderate or vigorous activities ranging from daily to 1-3 times per month, whereas our study required at least weekly MVPA, a higher threshold that may have amplified inverse associations.

Second, cultural context differs substantially; the United States has been described as “an outlier of religiosity in the developed world,” while Great Britain has undergone secularization leading to polarization between a larger secular group and smaller religiously fervent communities.^37–39^ Cross-sectional data from the Jackson Heart Study suggest that American religious communities actively promote health behaviours, which may not occur to the same extent in English religious settings.^40^ This suggests that religious attenders in our ELSA cohort may represent a fundamentally different population from those in American or broader European studies.

Third, covariates differed across studies: our analysis did not control for baseline functional limitations, which is important given that research shows instrumental functional limitations have the largest effect on changes in religious attendance among older adults.^41,42^ If individuals experiencing early functional decline increased religious engagement while reducing physical activity due to declining capacity, this could produce the inverse association we observed. Additionally, the dimensions of religiosity measured may interact with this reverse causation pathway: our finding that attendance, importance of faith, and religious purpose were all independently associated with less MVPA may reflect intensified religiosity as a coping response to declining health, rather than the stable “restful religiousness” (combining organizational involvement with religious education) that SHARE found to be health-promoting.^36,43^ The longer follow-up periods and multi-wave designs of the HRS and SHARE studies may have better controlled for these dynamic processes than our 4-year, two-wave design.

### R/S, Nutrition and HbA1c

Higher baseline non-organizational religiosity predicted future improvements in fruit and vegetable intake, independently of covariates. Our findings align with a systematic review of observational studies examining the association between R/S and fruit and vegetable intake.^10^ However, these studies were cross-sectional, and none evaluated non-organizational religiosity. We identified four additional cross-sectional studies published since this review that assessed R/S and fruit and vegetable intake; two reported a positive association,^44,45^ and two found no association.^46,47^ The only longitudinal study to date examined the ALSPAC cohort in Southwest England and found that religious attendance was associated with higher “health-conscious” dietary pattern scores, though it did not specifically assess fruit and vegetable intake or non-organizational religiosity.^48^ To our knowledge, this is the first study to use a longitudinal design to examine and report an independent association between higher non-organizational religiosity and increased future fruit and vegetable intake. One possible explanation is that, by improving eating awareness and reducing perceived stress, these practices may promote healthier dietary choices.^49^

Surprisingly, our results indicate that more regular attendance is prospectively associated with higher HbA1c levels, independent of covariates. Cross-sectional research on this relationship is limited and yields inconsistent findings, with most studies reporting no association between R/S and HbA1c or fasting plasma glucose in older adults, diabetic populations, and African American adults.^40,50–53^ Longitudinal evidence provides more relevant comparisons. Our findings align with a prospective analysis of elderly diabetic individuals from the HRS cohort, which indicated that religiosity predicted higher HbA1c levels over time.^17^ Similarly, in the Nurses’ Health Study II, the most frequent attenders had elevated risk of incident type 2 diabetes after full adjustment over 14 years.^54^ In contrast, the MIDUS study found no significant associations between multiple R/S measures and insulin resistance over 8-10 year.^55^ While the evidence remains mixed, findings from our ELSA cohort, the HRS cohort, and the Nurses’ Health Study II suggest that R/S may be associated with worse glycaemic outcomes in certain populations, warranting further investigation.

There are several possible explanations for this. For example, research has consistently shown that greater religiosity/spirituality is associated with obesity risk,^8,14,19^ which is linked to insulin resistance. However, as in our study, Walker et al. controlled for BMI in their analysis, suggesting that the effects of R/S on HbA1c may be independent of this factor.^17^ Furthermore, the dimension of R/S involved could reflect either a positive or a negative relationship. Specifically, the association was significant for regular attendance and, before adjustment, for the dimensions reflecting religious meaning and salience. However, a sense of purpose in life has been associated with better glycaemic control in observational studies,^56^ and attendance is the dimension most consistently linked to favourable health outcomes.^5^ Conversely, strongly held religious meaning may also reflect negative religious coping (NRC), the maladaptive use of religious beliefs or behaviours when facing life’s stressors,^57^ which can adversely affect physical health^44,58^ and might lead to higher HbA1c levels. Since interpreting life’s difficulties as ‘a punishment from God’ or ‘an act of the Devil’ is characteristic of NRC, individuals may attend more regularly in search of protection.

### R/S, Hypertension and Substance Use

Notably, some of our findings became clearer after adjusting for sociodemographic covariates (e.g., age, education, and sex), whereas others remained nonsignificant. First, consistent with previous evidence,^5,59^ all our R/S predictors protected against excessive alcohol intake. However, this association did not persist after controlling for covariates. Prior research on R/S and substance use disorders has suggested that NRC can reduce self-efficacy in managing substance use.^60^ Since men with alcohol use disorder are more likely to engage in these coping strategies,^61^ sex could have influenced our results. Second, DBP (but not SBP) was lower among those practising non-organizational R/S activities, such as praying or meditating daily and considering religion important, before adjustment. Although earlier literature linking hypertension and R/S has been somewhat inconsistent,^3^ meditation practices such as mindfulness have proved effective in lowering BP.^62–64^ Emerging longitudinal evidence suggests that R/S may protect against hypertension: the Nurses’ Health Study II found that religious attendance inversely associated with incident hypertension,^65^ whereas the Black Women’s Health Study found that R/S coping reduced hypertension risk, particularly under higher stress.^13^ The absence of association in model 3 might be due to age, education, or sex, as prior studies have shown lower BP among individuals with lower education,^66^ older age,^67^ and between women and men.^68^ Finally, contrary to both Koenig’s and Balboni’s reviews, we found no link between R/S and smoking status.^5,8^ However, longitudinal data from the MIDUS cohort showed that high religious involvement was associated with lower odds of being a persistent smoker but was not associated with smoking cessation among baseline smokers.^69^ This suggests R/S may be more protective against smoking initiation than cessation, which could explain our null findings in an older population where smoking patterns are already established.

### Strengths and Limitations

The analyses in this study have several strengths. Firstly, they are based on a large, representative sample of England’s older population. Secondly, they go beyond cross-sectional associations because of the prospective longitudinal design. Lastly, while previous studies mainly focused on one aspect of R/S, this study examines it as a multidimensional construct.

However, several limitations should be considered when interpreting our results. First, as with any prospective cohort study, unmeasured confounders that influence both R/S and cardiovascular risk factors, such as medication use, personality traits, social support, and well-being, could partly explain our findings.^4^ Future research would benefit from adjusting for these factors to avoid overestimating the impact of R/S on CVD risk. Second, multiple statistical tests were conducted, increasing the likelihood that some associations reached significance by chance alone. Across the 70 tests in our fully adjusted models (7 R/S measures × 10 outcomes), approximately 3.5 would be expected to be significant by chance at α=0.05; 9 were. This exceeds chance expectation, but by a modest margin, and our findings should therefore be read as suggestive rather than conclusive. Third, attrition may have affected the generalisability of this study’s outcomes. Lastly, generalisability is limited because most participants were White and Christian. Because religious traditions differ in the practices they emphasize, and practices may relate differently to cardiovascular risk, our findings may not extend to other traditions or to these as they shift over time.

## Conclusion

In conclusion, higher R/S showed mixed prospective associations with the cardiovascular risk profile. It was independently associated with lower fibrinogen, an inflammatory risk marker, and greater fruit and vegetable intake, but also with worse glycaemic control and lower participation in moderate-to-vigorous physical activity. Associations with DBP and alcohol consumption did not persist after adjustment, and we observed no prospective association with SBP, smoking status or CRP. Given the complexity of R/S, further research should include measures of NRC to clarify the mechanisms underlying these less favourable associations.

## Data Availability

The data analysed in this study are third-party data from the English Longitudinal Study of Ageing (ELSA) and were not generated by the authors. ELSA data are not openly available: access requires application to and approval from the ELSA study team, with approved users obtaining datasets through the UK Data Service (Study Number 5050). Further information on data access is available at https://www.elsa-project.ac.uk/accessing-elsa-data.

## Acknowledgements

We wish to thank all collaborators and participants in the ELSA Study for their valuable contributions. The authors used a large language model (Claude, Anthropic) to assist with manuscript editing, reference verification and preparation of the graphical abstract. All content was reviewed and verified by the authors, who take full responsibility for the work.

## Previous Presentation

Presented as a poster at the ESC Congress 2021 – The Digital Experience (27–30 August 2021) and published as an abstract in the European Heart Journal 2021;42(Suppl 1).

