## Supplemental Table for "Matters of the Heart and Soul: The Mixed Impact of Religiosity and Spirituality on the Cardiovascular Risk Profile Over Time"

### Supplemental Table 1. Variables and respective waves of collection

| Variables | Baseline (T0) | | Follow-up (T1) | |
|---|---|---|---|---|
| | Wave 4 (2008/09) | Wave 5 (2010/11) | Wave 6 (2012/13) | Wave 7 (2014/15) |
| **Covariates and Sociodemographic** | | | | |
| Age | | √ | | |
| Wealth | | √ | | |
| Education | | √ | | |
| Ethnicity | | √ | | |
| Gender | | √ | | |
| Religion category | | √ | | |
| Body-mass index | √ | | | |
| **Exposures** | | | | |
| Religiosity/Spirituality | | √ | | |
| Religious Attendance | | √ | | |
| Importance in everyday life | | √ | | |
| **Outcomes** | | | | |
| Systolic BP | √ | | √ | |
| Diastolic BP | √ | | √ | |
| CRP | √ | | √ | |
| Fibrinogen | √ | | √ | |
| HbA1c | √ | | √ | |
| Smoking status | | √ | | √ |
| Moderate/Intense PA | | √ | | √ |
| Sedentary Behaviour | | √ | | √ |
| Alcohol Units | | √ | | √ |
| Fru/Veg Intake | | √ | | √ |

√ = Collected; PA = Physical Activity; Fru/Veg = Fruits and Vegetables; CRP = C-Reactive Protein; BP = Blood Pressure; HbA1c = Glycated Haemoglobin.

### Supplemental Table 2. Baseline associations between religiosity/spirituality measures and sociodemographic characteristics, English Longitudinal Study of Ageing (ELSA), 2010/11

| | R/S scale score | | Importance of religion in daily life | | Religious attendance | |
|---|---|---|---|---|---|---|
| **Baseline characteristic** | **r** | **p** | **r** | **p** | **r** | **p** |
| Age group | .188 | <0.001 | .170 | <0.001 | .137 | <0.001 |
| Sex | .135 | <0.001 | .164 | <0.001 | .081 | <0.001 |
| Ethnicity | .122 | <0.001 | .147 | <0.001 | .128 | <0.001 |
| Wealth quintile | -.029 | 0.018 | -.035 | 0.004 | .047 | <0.001 |
| Education | -.030 | 0.014 | -.011 | 0.381 | .087 | <0.001 |

Values are Pearson correlation coefficients (r) with two-tailed p-values, computed on the analytic sample at baseline (n = 6,506–6,782 per pair; pairwise deletion).

Coding of baseline characteristics. Age group: 1 = 52–59, 2 = 60–69, 3 = 70–79, 4 = 80+. Sex: 1 = men, 2 = women. Ethnicity: 1 = White; 2 = Asian or Asian British, Black, African, Caribbean or Black British, Mixed ethnic group, or any other ethnic group (combined). Wealth: 1 = lowest quintile to 5 = highest. Education: 1 = no qualifications to 5 = degree.

Ethnic groups were combined because each comprised fewer than 80 participants (range 14–79), too few to support stable estimation; no comparisons between individual groups were made.

A positive coefficient indicates higher religiosity/spirituality at the higher code — that is, among older participants, among women, and among participants in the combined ethnic group category.
